# Burden and Economic Impact of Respiratory Viral Infections in Adults Aged 60 and Older: A Focus on RSV

**DOI:** 10.1101/2024.12.19.24319318

**Authors:** A. Peláez, S. Jimeno, M. Villarreal, M. Gil, I. Gutiérrez, M. Sanz, S. Natalini

## Abstract

**Background/Objectives:** Respiratory syncytial virus (RSV) represents a significant cause of acute respiratory infections (ARI) in adults aged 60 years and older, often leading to severe clinical outcomes and high healthcare costs. This study aimed to evaluate the clinical and economic burden of RSV compared to other ARI, focusing on specific age groups, comorbidities, and demographic factors.

**Methods:** A retrospective observational study was conducted using electronic medical records of adults aged ≥60 years hospitalized for ARI, including RSV, in Spain. Direct costs related to hospitalizations, intensive care unit (ICU) admissions, and treatments were analyzed. The study also assessed demographic, clinical, and comorbidity-related factors influencing the economic burden.

**Results:** RSV infections resulted in significantly higher direct costs compared to other ARI, particularly in patients aged 70–80 years. Comorbidities such as asthma and smoking history were associated with increased costs in RSV cases. Although ICU costs were comparable between groups, hospitalizations for RSV required longer stays and more intensive treatments, amplifying the overall economic burden. Differences in costs by age and sex highlighted the need for tailored clinical management strategies.

**Conclusions:** RSV poses a substantial economic and clinical burden on adults aged 60 years and older, particularly in those with comorbidities. Preventive measures, such as vaccination, could reduce healthcare costs and improve outcomes in this vulnerable population. These findings support the inclusion of RSV vaccines in immunization programs, especially in aging populations like Spain, to alleviate healthcare pressures during peak respiratory disease seasons.

## 1. Introduction

Respiratory infections represent one of the leading causes of morbidity and mortality globally, particularly among older adults, with a significant impact on public health systems and healthcare costs (1-3). In 2016, respiratory infections were responsible for approximately 2.38 million deaths worldwide, ranking as the sixth leading cause of overall mortality and the leading cause of death in children under five years old (4, 5). According to a recent study, the average cost of hospitalization for acute respiratory infection is notably higher in older patients and those with comorbidities, highlighting the growing economic burden this represents for healthcare systems (6). In addition, there is a significant increase in the population over 65 years of age in high-income countries, which makes the problem even more important (7).

The contribution of different viral agents in acute respiratory diseases in elderly patients has generated significant clinical interest, and thanks to the development of viral detection techniques, knowledge in this area has improved over the past 20 years (8, 9). However, diagnostic challenges persist due to difficulties in clinically recognizing the disease and the insufficient availability of routine tests and their limitations (8, 10-12).

Despite advancements in diagnostic techniques, RSV infection remains underdiagnosed, especially in patients with low viral load (13-16). The insufficient availability of rapid diagnostic tests and challenges in clinical recognition hinder the early identification and proper treatment of this infection in vulnerable populations, such as the elderly (1). RSV infection is a major concern in nursing homes due to the high transmission rates and increased susceptibility to respiratory illness in this population (17). In recent years, this viral agent has been linked to an elevated risk of severe complications in elderly individuals or those with chronic diseases (3, 18-23). RSV is responsible for a significant number of hospitalizations and deaths annually in adults, with estimates suggesting over 470,000 hospitalizations and 33,000 deaths in industrialized countries, underscoring the urgent need for more effective prevention and treatment strategies (24).

The economic impact of RSV on healthcare systems is considerable, with studies indicating that medical care related to RSV in older adults generates billions of dollars in annual cost, both in direct and indirect costs (25, 26). In Spain, hospitalizations related to RSV represent a significant expenditure for the National Health System (27, 28), emphasizing the need for measures to mitigate this economic impact and improve resource allocation in healthcare. The relationship between RSV and other viral respiratory infections, such as influenza and human metapneumovirus, further contributes to a scenario of high costs and healthcare service overload, particularly in the aging population (29, 30).

This situation highlights the urgent need to prioritize prevention and early diagnosis of respiratory infections in older adults (15, 31, 32). By focusing on improving vaccination and treatment strategies, as well as strengthening epidemiological surveillance, healthcare systems can optimize available resources and reduce the burden of these infections. The analysis of the economic burden of viral respiratory infections in older adults, particularly those with comorbidities, is essential for guiding decision-making in public health and ensuring that resources are used efficiently and effectively (6, 16). In this context, our study focuses on evaluating the economic impact of viral respiratory infections, specifically RSV, in adults aged 60 and over across different regions of Spain, with the aim of providing evidence to support the design of public policies and more effective intervention strategies.

## 2. Materials and Methods

A retrospective, multicenter observational study was conducted on older adult patients (≥60 years) diagnosed with RSV infection, confirmed by PCR or antigen testing, between October 1, 2023, and March 31, 2024. Inclusion criteria required hospitalization due to RSV infection confirmed by antigen or PCR tests.

Sociodemographic and clinical variables were collected, along with results from antigen and PCR tests for respiratory viruses. Data on pharmacological treatments, oxygen therapy requirements, and the need for invasive or non-invasive ventilation were also gathered. Information was extracted from the Doctoris electronic health record system, ensuring all patient identifiers were anonymized to protect confidentiality.

The study was conducted in accordance with the Declaration of Helsinki and approved by the Ethics Committee of HM Hospitales (protocol code 24.04.2344-GHM and date of approval: 8 May 2024) for studies involving humans. Moreover, this research is a retrospective study using anonymized data, which involves no direct patient intervention; therefore, individual consent to participate is not necessary.

Direct healthcare costs were estimated based on the type of care received. Average costs included €1457.60 per intensive care unit (ICU) stay, €744.16 for hospital admission, €198.91 per emergency room consultation, and €8.15 per patient for oxygen therapy. The cost of non-invasive ventilation (NIV) was €249.88, while invasive mechanical ventilation (IMV) was estimated at €312.34 per patient.

A preliminary descriptive analysis of patient characteristics was performed. Measures of central tendency and dispersion were calculated for quantitative variables, while counts and percentages were used for qualitative variables. Differences between groups were assessed using normality tests (Shapiro-Wilk, Kolmogorov-Smirnov) and homogeneity tests (Levene Test). Based on these results, parametric tests (ANOVA, Student’s t-test) or non-parametric tests (Kruskal-Wallis, Wilcoxon rank-sum) were applied. For qualitative variables, χ^2^ or Fisher’s exact tests were used.

To assess the impact of RSV vaccination, an efficacy value supported by previous studies was used. These projections indicated that vaccination could reduce severe RSV cases by 94.1% in the first year and 82.7% in the second year, whereas nonsevere cases could decrease by 82.6% and 74.5%, respectively (33). This efficacy value was used to estimate potential reductions in hospitalizations and other RSV-related outcomes in the study population.

To identify and quantify comorbidities with significant impacts on healthcare costs, mixed-effects models were developed. These included a simple linear model and mixed-effects models with random intercept, random slope, and both random intercept and slope. Model comparisons were based on their fit to the logarithm of total cost per patient. A random sample of patients with high-cost comorbidities was used for case reduction estimations. Data analysis and visualization were conducted using RStudio.

## 3. Results

### 3.1. Socio-demographic, Clinical, and Healthcare Costs Comparison Between Respiratory Diseases and RSV

Total costs associated with episodes of acute respiratory infections (ARI) and respiratory syncytial virus (RSV) cases were compared across various socio-demographic and clinical aspects (Table 1). The data show that the average total cost for ARI episodes is €3,329.17 (±€7,175.97), while for RSV episodes it is €5,196.96 (±€10,019.56), with a statistically significant difference (p < 0.001).

**Table 1.** Total cost for each socio demographic and clinical characteristics for ARI in comparison with RSV cases.

| Characteristics | Total ARI<br>(n1 =1,952) | Total RSV cases<br>(n2= 229) | p-value |
| --- | --- | --- | --- |
| **No. of re-admissions** |  |  |  |
| 1 admission | 3,588.41 (±7,502.95) | 8,075.52 (±13,940.15) | <0.001 |
| 2 admissions | 3,017.18 (±6,875.64) | 3,416.96 (±4,540.46) | <0.001 |
| >2 admissions | 3,095.21 (±6,445.11) | 2,721.14 (±4,156.28) | <0.001 |
| **Sex [Female]** |  |  |  |
| Male | 3,751.76 (±7,889.96) | 6,884.91 (±14,269.76) | <0.001 |
| Female | 2,972.09 (±6,494.35) | 4,000.28 (±4,970.47) | <0.001 |
| **Age group** |  |  |  |
| 60-70 years old | 2,975.96 (±7,849.17) | 3,980.7 (±4,237.2) | <0.001 |
| 70-80 years old | 3,370.93 (±8,510.16) | 7,800.36 (±17,474.31) | <0.001 |
| 80-90 years old | 3,421.95 (±5,586.12) | 4,319.5 (±6,285.38) | 0.140 |
| +90 years old | 3,716.71 (±4,752.43) | 4,891.5 (±5,692.02) | 0.170 |
| **Days of hospitalisation** |  |  |  |
| <10 days | 3,606.67 (±1,806.36) | 3,545.38 (±1,956.24) | 0.981 |
| 10-30 days | 1,1695 (±6,052.03) | 9,830.16 (±5,561.21) | 0.117 |
| >30 days | 43,620.57 (±22,863.61) | 52,628.73 (±39,088.03) | 0.813 |
| **UCI** | 21,006.79 (±22,851.57) | 21,828.7 (±33,100) | 0.773 |
| **Days of ICU hospitalisation** |  |  |  |
| <10 days | 18,070.09 (±20,022.44) | 21,828.7 (±33,100) | 0.975 |
| 10-30 days | 45,105.77 (±13,636.62) | NA (±NA) | NA |
| >30 days | 100,813.28 (±NA) | NA (±NA) | NA |
| **Exitus** | 12,661.34 (±13,701.38) | 8,042.49 (±6,768.67) | 0.454 |
| **Neoplasia** | 4,908.82 (±9,731.23) | 7,449.31 (±15,737.77) |  |
| **Asthma** | 3,169.43 (±4,064.99) | 5,793.11 (±4,880.71) | 0.035 |
| **Current smoker** | 5,099.17 (±8,507.84) | 5,109.19 (±6,316.44) | 0.502 |
| **Former smoker** | 4,408.68 (±6,855.51) | 5,736.77 (±6,893.49) | 0.021 |
| **Alcoholism** | 7,124.05 (±16,753.31) | 21,007.81 (±43,829.38) | 0.156 |
| **Diabetes** | 4,454.19 (±7,891.93) | 5,031.63 (±5,194.22) | 0.078 |
| **Obesidad** | 3,872.33 (±5,250.17) | 3,983.81 (±3,732.9) | 0.422 |
| **Hipertensión** | 3,969.77 (±7,317.09) | 5,113.65 (±11,124.21) | 0.055 |
| **EPOC** | 4,835.97 (±8,535.16) | 5,020.58 (±5,617.36) | 0.255 |
| **Insuficiencia renal** | 6,288.67 (±11,776.8) | 8,961.8 (±17,227.29) | 0.127 |
| **Insuficiencia renal cardiaca** | 5,136.56 (±8,545.83) | 6,574.83 (±14,927.49) | 0.566 |
| **CCI** |  |  |  |
| 0 | 1,480.46 (±4,646.95) | 3,538.16 (±6,809.92) | 0.011 |
| 1 | 3,214.08 (±4,602.83) | 5,108.24 (±5,134.96) | 0.046 |
| 2 | 4,108.37 (±7,822.67) | 3,902.92 (±3,645.29) | 0.158 |
| 3 | 5,399.81 (±13,193.51) | 8,495.83 (±25,791.21) | 0.810 |
| 4 | 4,591.78 (±6,976.44) | 5,410.87 (±6,587.38) | 0.261 |
| ≥5 | 4,936.1 (±8,448.64) | 5,689.64 (±7,801.82) | 0.160 |
| **Community** |  |  |  |
| Andalucía | 2,289.25 (±3,978.57) | NA (±NA) | NA |
| Barcelona | 3,236.12 (±7,884.45) | 2,494.36 (±3,614.44) | 0.891 |
| Galicia | 2,080.4 (±3,809.47) | 2,685.84 (±3,564.29) | 0.218 |
| León | 925.79 (±1,711.17) | NA (±NA) | NA |
| Madrid | 3491.1 (±7,313.29) | 5,528.96 (±10,526.6) | **<0.001** |
| **Total cost** | 3,329.17 (±7,175.97) | 5,196.96 (±10,019.56) | **<0.001** |

ta are shown as n (%), mean ±sd. Significant comparisons (p < 0.05) are marked in bold.

In terms of costs related to readmissions, patients with RSV incur significantly higher cost compared to ARI patients. For a single admission, the average cost in RSV cases is €8,075.52 (±€13,940.15), compared to €3,588.41 (±€7,502.95) in ARI (p < 0.001). For two readmissions, the average costs are €3,416.96 (±€4,540.46) for RSV and €3,017.18 (±€6,875.64) for ARI (p < 0.001). For more than two readmissions, costs are €2,721.14 (±€4,156.28) for RSV versus €3,095.21 (±€6,445.11) for ARI (p < 0.001).

Regarding age groups, patients aged 70-80 years with RSV incur significantly higher costs (€7,800.36 ±€17,474.31) compared to ARI patients (€3,370.93 ±€8,510.16) (p < 0.001). However, for patients aged 80-90 years and over 90 years, the cost differences between RSV and ARI are not statistically significant (p = 0.140 and p = 0.170, respectively).

Total costs associated with ICU hospitalization show that the average cost is €21,006.79 (±€22,851.57) for ARI and €21,828.70 (±€33,100.00) for RSV, with no significant differences between the two groups (p = 0.773).

Regarding comorbidities, patients with RSV have higher costs compared to ARI patients for asthma (€5,793.11 ±€4,880.71 vs €3,169.43 ±€4,064.99, p = 0.035) and for former smokers (€5,736.77 ±€6,893.49 vs €4,408.68 ±€6,855.51, p = 0.021). However, the cost differences for other comorbidities analyzed, such as diabetes, hypertension, and COPD, are not statistically significant.

### 3.2. Total Costs and RSV Case Reduction

A cost analysis of RSV cases was conducted from October 2023 to March 2024 (Table 2; Figure 1) to assess the economic impact of introducing the RSV vaccine. The study projected that the vaccine would reduce severe RSV cases by 94.1% in the first year and by 82.7% in the second year of protection. In non-severe cases, reductions were expected to be 82.6% in the first year and 74.5% in the second year.

**Table 2.** Estimate and expected number of cases of RSV and total cost before and after vaccine application.

| RSV month | Period | VRS Cases | Reduced cases due to vaccine<sup>1</sup> | Total cost (€) of sanitary uses | Total cost (€) of sanitary uses after using vaccine | % of change |
| --- | --- | --- | --- | --- | --- | --- |
| 10-2023 | 1 year | 5 | 0 | 25,699.26 | 0.0 | -100.0 |
|  | 2 year |  | 0 |  | 0.0 | -100.0 |
| 11-2023 | 1 year | 46 | 4 | 402,720.967 | 13,991.61 | -96.5 |
|  | 2 year |  | 9 |  | 54,543.35 | -86.5 |
| 12-2023 | 1 year | 77 | 6 | 392,023.8 | 13,991.61 | -96.4 |
|  | 2 year |  | 16 |  | 73,575.89 | -81.2 |
| 01-2024 | 1 year | 70 | 8 | 252,768.07 | 28,528.47 | -88.7 |
|  | 2 year |  | 15 |  | 41,001.04 | -83.8 |
| 02-2024 | 1 year | 10 | 0 | 56,062.39 | 0.0 | -100.0 |
|  | 2 year |  | 2 |  | 397.82 | -99.3 |
| 03-2024 | 1 year | 21 | 3 | 60,829.22 | 596.73 | -99.0 |
|  | 2 year |  | 5 |  | 12,702.2 | -79.1 |
| Total | 1 year | 229 | 21 | 1,190,103.71 | 89,448.42 | -92.5 |
|  | 2 year |  | 47 |  | 216,320.3 | -81.8 |

**Figure 1.**
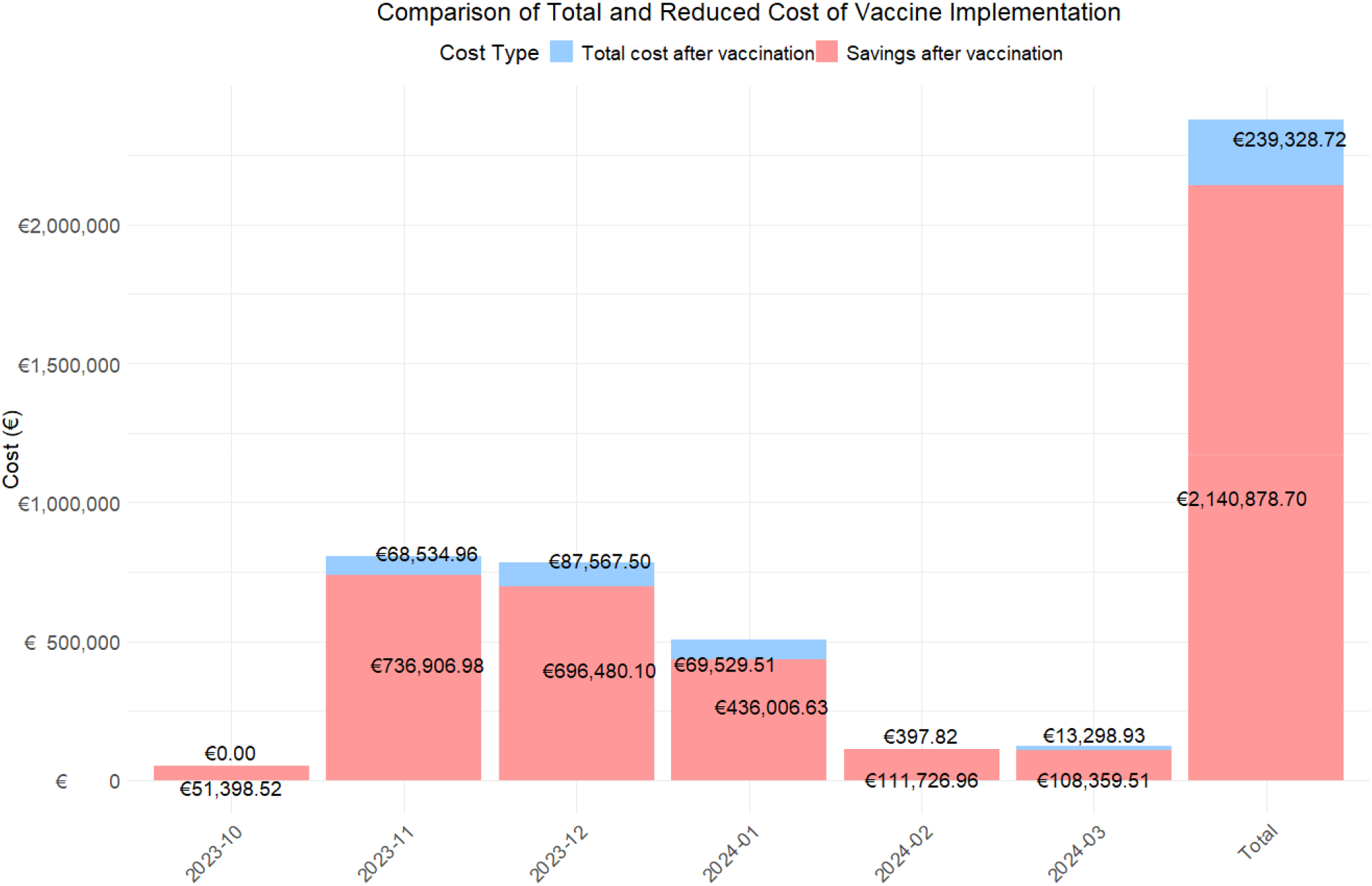
Bar chart to assess total expenditure and savings from vaccination using 1^st^ and 2^nd^ year cost.

The data show a notable reduction in monthly and total costs. The global analysis for the period from October 2022 to March 2023 revealed that the total cost associated with RSV cases without the vaccine was €1,190,103.71 from a total of 229 RSV cases. With the introduction of the hypothetical vaccine, cases would be reduced to 21 in the first year and 47 in the second year, with an associated cost of €89,448.42 and €216,320.30, representing an overall decrease of 92.5% and 81.8%.

When analyzed by month, the three months with the highest costs during the RSV season were November (€402,720.97), December (€392,023.80), and January (€252,768.07), coinciding with the epidemic peak. During these months, 46 cases, 77 cases, and 70 RSV cases were reported, respectively. With the introduction of the hypothetical vaccine, it is projected that RSV cases will be reduced to 13, 22 and, 23 cases in two years, respectively. This would result in total costs of €13,991.61 and €54,543.35, €13,991.61 and €73,575.89, and €28,528.47 and €41,001.04, representing cost reductions of 96.5% and 86.5%, 96.4% and 81.2%, and 88.7% and 83.8% in each of these months, respectively.

When analyzing the costs associated with the use of the vaccine over two years (Figure 1), the total cost after its implementation is estimated at €239,328.72, resulting in savings of €2,140,878.70 over this period. Furthermore, during the peak epidemic months, the estimated savings would amount to €68,534.96, €87,567.50, and €69,529.51, respectively.

### 3.3. Total Costs and RSV Case Reduction by age range

To more thoroughly assess the financial impact of the RSV vaccine, a detailed analysis was conducted across different age groups, considering both the estimated number of patients and the total healthcare costs before and after vaccination (Table 3; Figures 2 and 3).

**Table 3.**
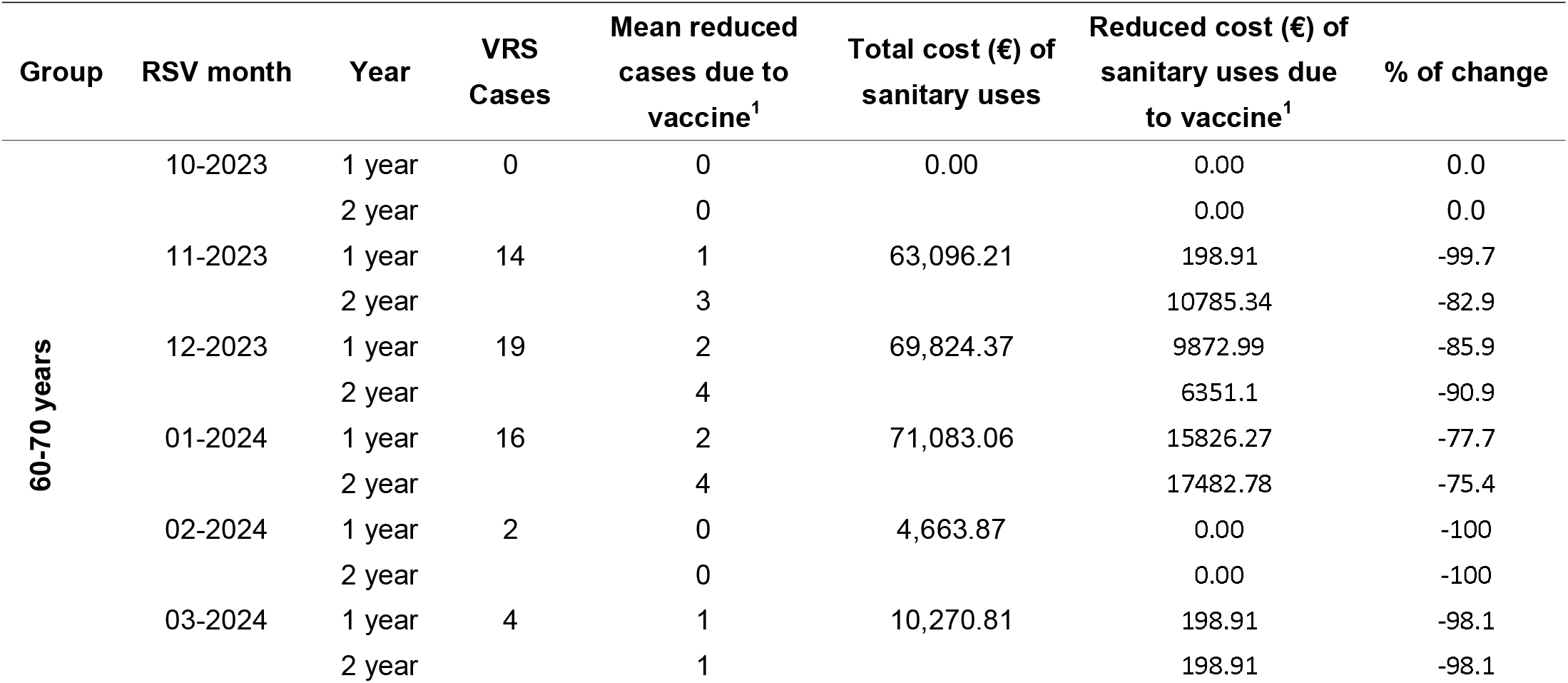
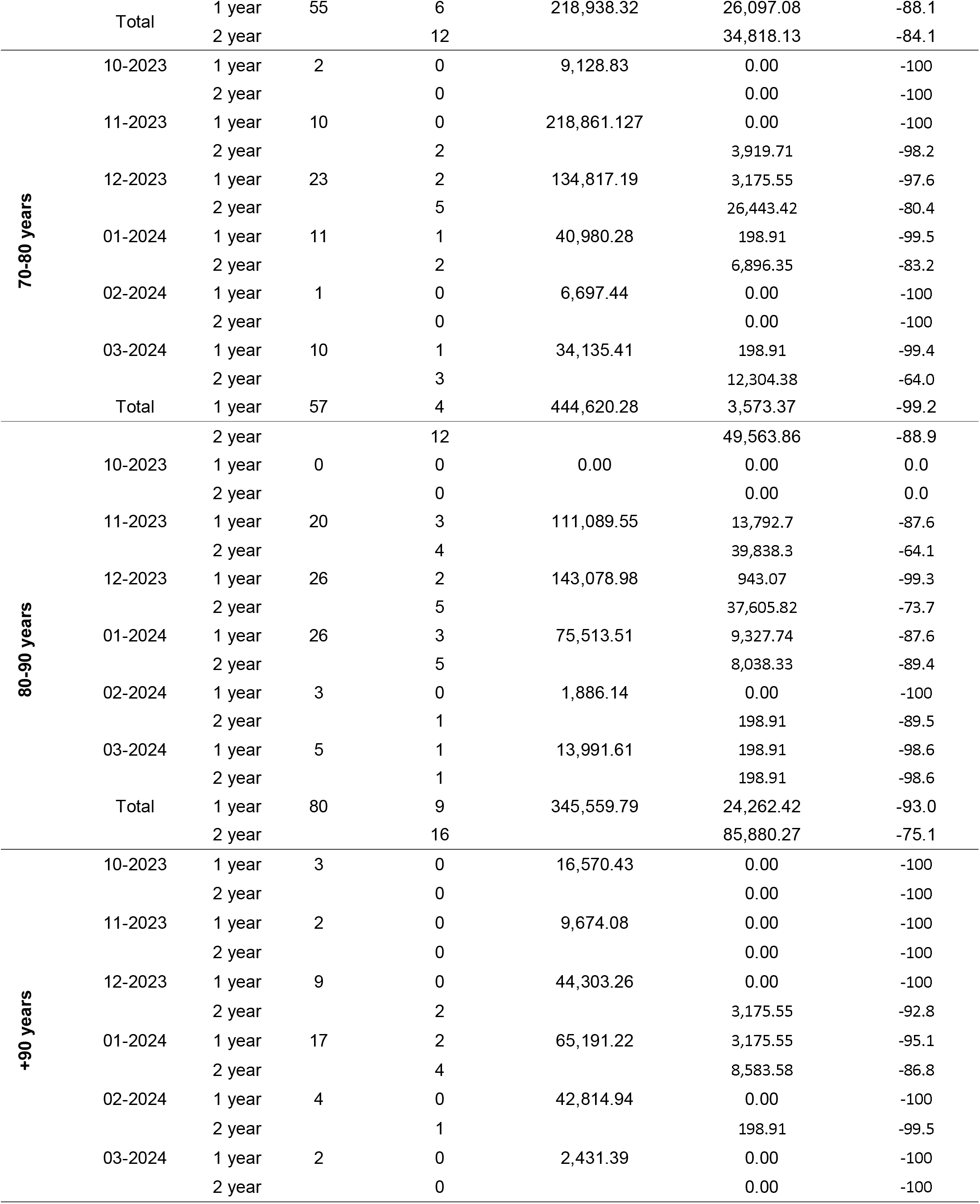
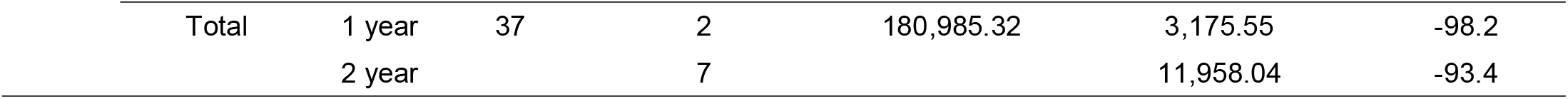
Estimated and expected number of patients and total cost before and after vaccine implementation, segregated by age groups.

| Group | RSV month | Year | VRS Cases | Mean reduced cases due to vaccine<sup>1</sup> | Total cost (€) of sanitary uses | Reduced cost (€) of sanitary uses due to vaccine<sup>1</sup> | % of change |
| --- | --- | --- | --- | --- | --- | --- | --- |
| 60-70 years | 10-2023 | 1 year | 0 | 0 | 0.00 | 0.00 | 0.0 |
|  |  | 2 year |  | 0 |  | 0.00 | 0.0 |
|  | 11-2023 | 1 year | 14 | 1 | 63,096.21 | 198.91 | -99.7 |
|  |  | 2 year |  | 3 |  | 10785.34 | -82.9 |
|  | 12-2023 | 1 year | 19 | 2 | 69,824.37 | 9872.99 | -85.9 |
|  |  | 2 year |  | 4 |  | 6351.1 | -90.9 |
|  | 01-2024 | 1 year | 16 | 2 | 71,083.06 | 15826.27 | -77.7 |
|  |  | 2 year |  | 4 |  | 17482.78 | -75.4 |
|  | 02-2024 | 1 year | 2 | 0 | 4,663.87 | 0.00 | -100 |
|  |  | 2 year |  | 0 |  | 0.00 | -100 |
|  | 03-2024 | 1 year | 4 | 1 | 10,270.81 | 198.91 | -98.1 |
|  |  | 2 year |  | 1 |  | 198.91 | -98.1 |
|  | Total | 1 year | 55 | 6 | 218,938.32 | 26,097.08 | -88.1 |
|  |  | 2 year |  | 12 |  | 34,818.13 | -84.1 |
| 70-80 years | 10-2023 | 1 year | 2 | 0 | 9,128.83 | 0.00 | -100 |
|  |  | 2 year |  | 0 |  | 0.00 | -100 |
|  | 11-2023 | 1 year | 10 | 0 | 218,861.127 | 0.00 | -100 |
|  |  | 2 year |  | 2 |  | 3,919.71 | -98.2 |
|  | 12-2023 | 1 year | 23 | 2 | 134,817.19 | 3,175.55 | -97.6 |
|  |  | 2 year |  | 5 |  | 26,443.42 | -80.4 |
|  | 01-2024 | 1 year | 11 | 1 | 40,980.28 | 198.91 | -99.5 |
|  |  | 2 year |  | 2 |  | 6,896.35 | -83.2 |
|  | 02-2024 | 1 year | 1 | 0 | 6,697.44 | 0.00 | -100 |
|  |  | 2 year |  | 0 |  | 0.00 | -100 |
|  | 03-2024 | 1 year | 10 | 1 | 34,135.41 | 198.91 | -99.4 |
|  |  | 2 year |  | 3 |  | 12,304.38 | -64.0 |
|  | Total | 1 year | 57 | 4 | 444,620.28 | 3,573.37 | -99.2 |
|  |  | 2 year |  | 12 |  | 49,563.86 | -88.9 |
| 80-90 years | 10-2023 | 1 year | 0 | 0 | 0.00 | 0.00 | 0.0 |
|  |  | 2 year |  | 0 |  | 0.00 | 0.0 |
|  | 11-2023 | 1 year | 20 | 3 | 111,089.55 | 13,792.7 | -87.6 |
|  |  | 2 year |  | 4 |  | 39,838.3 | -64.1 |
|  | 12-2023 | 1 year | 26 | 2 | 143,078.98 | 943.07 | -99.3 |
|  |  | 2 year |  | 5 |  | 37,605.82 | -73.7 |
|  | 01-2024 | 1 year | 26 | 3 | 75,513.51 | 9,327.74 | -87.6 |
|  |  | 2 year |  | 5 |  | 8,038.33 | -89.4 |
|  | 02-2024 | 1 year | 3 | 0 | 1,886.14 | 0.00 | -100 |
|  |  | 2 year |  | 1 |  | 198.91 | -89.5 |
|  | 03-2024 | 1 year | 5 | 1 | 13,991.61 | 198.91 | -98.6 |
|  |  | 2 year |  | 1 |  | 198.91 | -98.6 |
|  | Total | 1 year | 80 | 9 | 345,559.79 | 24,262.42 | -93.0 |
|  |  | 2 year |  | 16 |  | 85,880.27 | -75.1 |
| +90 years | 10-2023 | 1 year | 3 | 0 | 16,570.43 | 0.00 | -100 |
|  |  | 2 year |  | 0 |  | 0.00 | -100 |
|  | 11-2023 | 1 year | 2 | 0 | 9,674.08 | 0.00 | -100 |
|  |  | 2 year |  | 0 |  | 0.00 | -100 |
|  | 12-2023 | 1 year | 9 | 0 | 44,303.26 | 0.00 | -100 |
|  |  | 2 year |  | 2 |  | 3,175.55 | -92.8 |
|  | 01-2024 | 1 year | 17 | 2 | 65,191.22 | 3,175.55 | -95.1 |
|  |  | 2 year |  | 4 |  | 8,583.58 | -86.8 |
|  | 02-2024 | 1 year | 4 | 0 | 42,814.94 | 0.00 | -100 |
|  |  | 2 year |  | 1 |  | 198.91 | -99.5 |
|  | 03-2024 | 1 year | 2 | 0 | 2,431.39 | 0.00 | -100 |
|  |  | 2 year |  | 0 |  | 0.00 | -100 |

|  |  |  |  |  |  |  |
| --- | --- | --- | --- | --- | --- | --- |
| Total | 1 year | 37 | 2 | 180,985.32 | 3,175.55 | -98.2 |
|  | 2 year |  | 7 |  | 11,958.04 | -93.4 |

**Figure 2.**
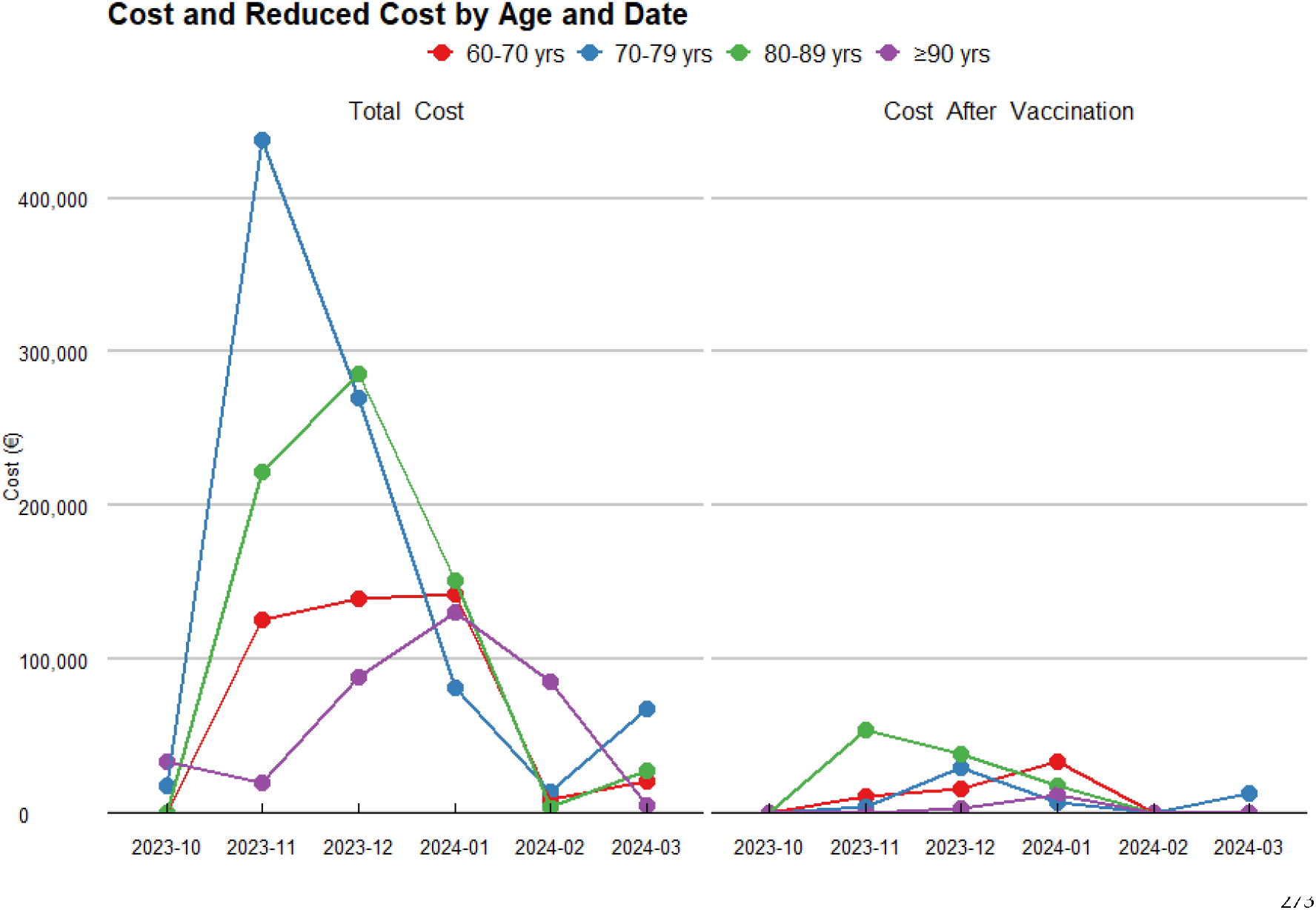
Temporal Evolution of Costs and their Reduction by Age Category.

**Figure 3.**
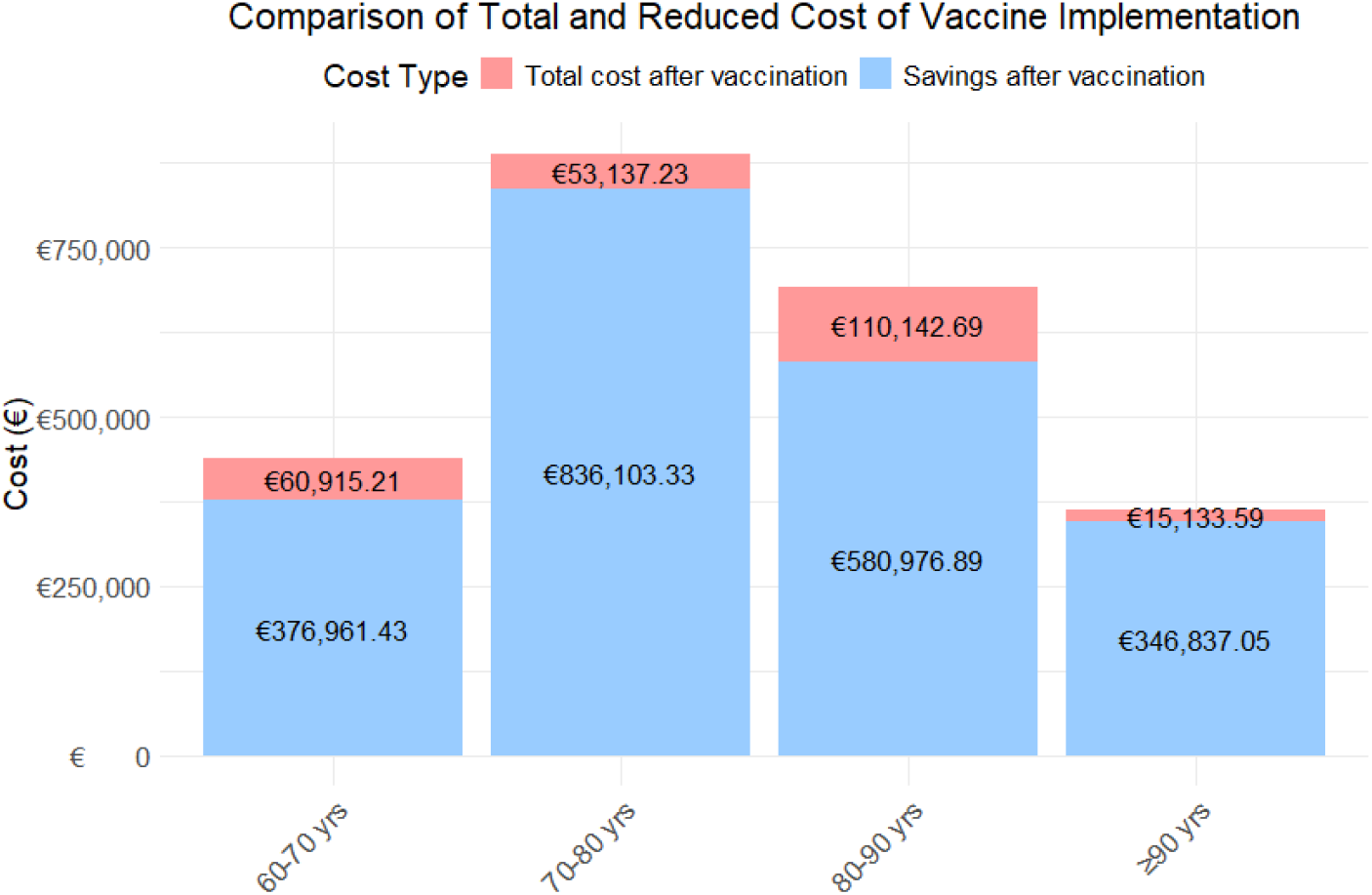
Costs and their Reduction by Age Category.

For the 60–70-year-old age group, the analysis projects that the vaccine will result in a substantial reduction in RSV cases, decreasing from 55 to 18 cases. This reduction leads to an 88.1% decrease in the first year, and 84.1% decrease in the second year in healthcare costs, dropping from an estimated total of €218,938.32 to €26,097.08 and €34,818.13 in the first and second year, respectively.

In the 70–80-year-old group, the vaccine is projected to reduce RSV cases from 57 to 16, reflecting a 99.2% decrease in the first year, and 88.9% decrease in the second year in healthcare costs, with savings of €444,620.28, reducing the total to €35,753.37 in the first year and €49,563.86 in the second year.

For the 80–90-year-old group, a significant reduction in RSV cases is observed, from 80 to 25, resulting in a 93.0% decrease in the first year, and 75.1% decrease in the second year. This reduction leads to savings of €345,559.79, bringing the total cost down to €24,262.42 and €85,880.27.

Finally, in the 90+ years age group, the vaccine has a notable impact, reducing RSV cases from 37 to 9 and cutting the associated healthcare costs by 98.2% decrease in the first year and 93.4% decrease in the second year. The total spending decreases from €180,985.32 to €3,175.55 and €11,958.04.

When analyzing the total costs by age group over the two-year period (Figure 3), the age group with the highest expenditure is 80–90 years, followed by 60–70 years, 70–80 years, and finally those aged ≥90 years. In contrast, the age group achieving the greatest savings is 70–80 years, followed by 80–90 years, and 90+ years age group.

### 3.4. Total Costs and RSV Case Reduction Due to the Most Important Comorbidities

In order to identify and quantify comorbidities that have a significant impact on total healthcare costs in a patient population, several mixed-effects models with varying complexity were developed. First, a simple linear model was created, along with a mixed-effects linear model with random intercept (patient identifier as a random effect to model individual variability), random slope (age as a random effect to capture heterogeneity between patients), and both random intercept and slope (identifier and age as random effects). The four models were compared to fit the logarithm of the total cost per patient. The simple linear model had the lowest AIC (6754.621) and BIC (6849.309), indicating the best balance between fit and simplicity. The mixed models with random intercept (AIC: 6800.141), random slope (AIC: 6808.580), and both random intercept and slope (AIC: 6812.580) did not show significant improvements in fit, according to the likelihood ratio test. Although the mixed models capture individual variability, the simple linear model proved to be the most suitable due to its simplicity and robust fit.

The results from the simple linear model (e.Table1) revealed significant associations between various comorbidities and healthcare costs. First, all evaluated comorbidities, except for obesity (coefficient = 0.051, p = 0.639), alcoholism (coefficient = 0.105, p = 0.518), and being a former smoker (coefficient = 0.093, p = 0.255), showed significant increases in total costs. Among the comorbidities that contributed most to the increase in healthcare spending, pneumonia and bronchitis were found, with coefficients of 1.942 and 1.783, respectively (p-values < 0.001). Additionally, acute infections showed higher increases in total cost (coefficients 1.412, p-value < 0.001), compared to the other significant variables. Using the results, we evaluated the total cost and potential reductions in healthcare expenditures that an RSV vaccine could bring for patients with comorbidities significantly impacting healthcare costs (Table 4).

**Table 4.** Impact of RSV Vaccine on Healthcare Costs and Case Reductions for Patients with Comorbidities.

| Comorbidity | Year | VRS Cases | Reduced cases due to vaccine<sup>1</sup> | Total cost (€) of sanitary uses | Reduced cost (€) of sanitary uses due to vaccine<sup>1</sup> | % of change |
| --- | --- | --- | --- | --- | --- | --- |
| Pneumonia | 1 year | 50 | 2 | 303,242.08 | 22,324.8 | -94.5 |
|  | 2 year |  | 8 |  | 36,463.84 | -91.0 |
| Bronchitis | 1 year | 29 | 2 | 192,672.72 | 3,720.8 | -98.1 |
|  | 2 year |  | 4 |  | 49,858.72 | -74.1 |
| Acute infections | 1 year | 23 | 1 | 130,866.76 | 198.91 | -99.8 |
|  | 2 year |  | 3 |  | 12,304.38 | -84.8 |
| Renal insufficiency | 1 year | 62 | 9 | 555,631.417 | 33,192.34 | -95.6 |
|  | 2 year |  | 11 |  | 60,496.63 | -92.0 |
| Current smoker | 1 year | 41 | 3 | 209,476.9 | 3,374.46 | -98.4 |
|  | 2 year |  | 8 |  | 29,073.72 | -86.1 |
| Heart failure | 1 year | 77 | 6 | 506,262.167 | 22,177.37 | -95.9 |
|  | 2 year |  | 11 |  | 17,763.89 | -96.7 |
| Asthma | 1 year | 14 | 1 | 81,103.48 | 9,674.08 | -96.81 |
|  | 2 year |  | 3 |  | 5,606.94 | -98.2 |
| Neoplasia | 1 year | 72 | 7 | 536,350.357 | 26,295.99 | -88.9 |
|  | 2 year |  | 15 |  | 73,973.71 | -68.7 |
| Hypertension | 1 year | 147 | 14 | 751,706.697 | 28,034.7 | -94.5 |
|  | 2 year |  | 30 |  | 104,525.79 | -79.4 |
| Diabetes | 1 year | 47 | 6 | 236,486.84 | 25,898.17 | -80.2 |
|  | 2 year |  | 5 |  | 17,482.78 | -86.6 |
| COPD | 1 year | 81 | 6 | 406,666.9 | 10,668.63 | -98.1 |
|  | 2 year |  | 15 |  | 68,366.77 | -87.7 |

## 4. Discussion

The findings of this study highlight the significant clinical and economic burden associated with RSV infections in adults aged 60 years and older. Compared to other ARIs, episodes caused by RSV result in substantially higher costs, particularly in specific age groups, such as patients aged 70–80 years. This underscores the importance of prioritizing prevention and control strategies for this vulnerable group, given their increased susceptibility to severe complications and associated hospital costs (3, 19, 21, 22).

The analysis of direct costs reveals that RSV-linked hospitalizations and readmissions represent a substantial economic burden on healthcare systems, significantly exceeding the costs observed for other ARIs. This increase is due to the need for intensive care, prolonged hospital stays, and more complex treatments, especially in patients with comorbidities. Furthermore, the significant differences observed among sex and age groups suggest that clinical management should consider demographic and clinical characteristics to optimize resource utilization.

Regarding comorbidities, conditions such as asthma or a smoking history are associated with higher costs in patients with RSV compared to those with other ARIs. This finding highlights the importance of identifying and managing risk factors that may exacerbate the clinical and economic impact of RSV (20, 23). Although no significant differences in the costs of ICU admissions were observed between groups, the overall ICU burden on healthcare systems remains notable and deserves attention.

From a public health perspective, these results underscore the value of preventive strategies, particularly RSV vaccination. Implementing immunization programs targeting older adults could yield benefits not only in health outcomes but also in economic savings (15, 31, 32). Vaccination could reduce immediate costs related to hospital care and complications, while improving the quality of life of patients with comorbidities (6, 16, 34). It should be noted that the effect of vaccination in these patients is especially valuable considering that the duration of protection offered by the vaccine extends beyond a single season, as demonstrated by several recent publications (33, 35). In addition, these vaccination programs for the most vulnerable population groups could reduce pressure on healthcare systems during peak respiratory disease seasons, optimizing resource allocation and reducing morbidity and mortality rates.

However, some limitations should be acknowledged when interpreting these results. The retrospective nature of the study and the reliance on electronic medical records could introduce biases in data collection. In addition, the analysis has focused exclusively on direct clinical costs, without considering indirect costs, such as loss of productivity or economic burden for caregivers. Future studies should address these gaps to provide a more comprehensive assessment of the economic and social impact of RSV in this population.

In conclusion, this study reinforces the need to prioritize preventive strategies against RSV, particularly vaccination, as a key tool to mitigate its clinical and economic burden in older adults. These findings provide strong evidence to support in routine national immunization programs, especially in countries with aging populations, such as Spain.

## 5. Conclusions

In conclusion, this study underscores the urgent need to prioritize preventive strategies against RSV, particularly vaccination, as a key tool to mitigate its clinical and economic burden in older adults.

## Supporting information

e.Table 1. Results obtained from the Simple Linear Model for Comorbidities Associated with Total Health Costs

## Author Contributions

Conceptualization, Jimeno S. and Natalini Martínez S.; methodology, Peláez A., Jimeno S. and Natalini Martínez S.; software, Peláez A.; validation, Jimeno S. and Natalini Martínez S.; formal analysis, Peláez A.; investigation, Peláez A., Jimeno S. and Natalini Martínez S.; resources, Peláez A.; data curation, Peláez A.; writing—original draft preparation, all authors; writing—review and editing, all authors; visualization, Peláez A.; supervision, Jimeno S. and Natalini Martínez S.; project administration, Natalini Martínez S.; funding acquisition, no funding. All authors have read and agreed to the published version of the manuscript.

## Institutional Review Board Statement

The study was conducted in accordance with the Declaration of Helsinki and approved by the Ethics Committee of HM Hospitales (protocol code 24.04.2344-GHM and date of approval: 2024/05/08) for studies involving humans.

## Informed Consent Statement

Informed consent was waived due to the retrospective nature of the study and the fact that no personal identifiable information was used.

## Data Availability Statement

The data supporting the reported results cannot be shared due to ethical restrictions.

