## Supplementary material for "Burden and Economic Impact of Respiratory Viral Infections in Adults Aged 60 and Older: A Focus on RSV": e.Table 1. Results obtained from the Simple Linear Model for Comorbidities Associated with Total Health Costs

| **Variable** | **Coefficients** | **Standard Errors** | **p-values** |
| --- | --- | --- | --- |
| **Intercept** | 5.855 | 0.051 | **<0.001** |
| **Pneumonia** | 1.942 | 0.098 | **<0.001** |
| **Bronchitis** | 1.783 | 0.143 | **<0.001** |
| **Acute infections** | 1.412 | 0.171 | **<0.001** |
| **Renal insufficiency** | 0.438 | 0.086 | **<0.001** |
| **Current smoker** | 0.323 | 0.107 | **0.002** |
| **Heart failure** | 0.297 | 0.085 | **<0.001** |
| **Asthma** | 0.272 | 0.138 | **0.049** |
| **Neoplasia** | 0.227 | 0.076 | **0.003** |
| **Hypertension** | 0.225 | 0.07 | **0.001** |
| **Diabetes** | 0.187 | 0.085 | **0.028** |
| **COPD** | 0.176 | 0.086 | **0.042** |
| **Alcoholism** | 0.105 | 0.162 | 0.518 |
| **Former smoker** | 0.093 | 0.082 | 0.255 |
| **Obesity** | 0.051 | 0.108 | 0.639 |

Variables with a significant association (p-value < 0.05) are marked in bold and with an asterisk.
